# Patterns of non-fatal overdose and injection-related bacterial infections during pregnancy and the postpartum year among New York State residents

**DOI:** 10.1101/2025.01.21.25320879

**Authors:** Hannah LF Cooper, Rohan R. D’Souza, Howard H. Chang, Emily Peterson, Erin Rogers, Simone Wien, Sarah C. Blake, Michael R. Kramer

**Affiliations:** Rollins School of Public Health, Emory University, 1518 Clifton Road NE, Atlanta, GA 30322

**Keywords:** Overdose, endocarditis, abscess, maternal, pregnancy, postpartum

## Abstract

**Objectives:** Overdoses are a leading cause of maternal mortality in the US, but limited evidence exists about patterns of nonfatal overdose, a key risk factor for subsequent fatal overdose, or of other drug-related harms. Here, we estimate prevalences of nonfatal overdose and injection-related endocarditis and abscesses/cellulitis across the 21 months spanning pregnancy and the postpartum year.

**Methods:** Among people who experienced an in-hospital birth in New York State between 9/1/2016 and 1/1/2018 (N=330,872), we estimated the prevalences of hospital-based diagnoses of nonfatal overdose and of injection-related bacterial infections (i.e., endocarditis, abscesses, and cellulitis) across these 21 months; by trimester and postpartum quarter; and by social position (e.g., race/ethnicity, rurality, payor).

**Results:** The 21-month nonfatal overdose prevalence was 158/100,000 births (CI: 145/100,000, 172/100,000); the 21-month prevalence of injection-related bacterial infections was 56/100,000 births (CI: 49/100,000, 65/100,000). There was a trend such that rates of overdose and of injection-related bacterial infections declined as pregnancy progressed and rebounded postpartum. Rates of all outcomes were highest outside of large metropolitan areas and among publicly insured residents.

**Conclusions for Practice:** The trend toward diminished rates during pregnancy is supported by past qualitative studies. If confirmed by future research in other geographical regions and with larger sample sizes, this finding holds promise for programmatic and policy interventions. Interventions co-designed with people who use drugs could complement and support harm reduction efforts that pregnant people are already engaging in independently. Such efforts can help people who use drugs survive the pregnancy and postpartum year.

**Significance:** “What is already known on this subject?

Fatal overdoses are a leading cause of maternal mortality in the US. Little evidence exists, however, about patterns of nonfatal overdose, a strong predictor of future fatal overdose, or about other serious injection-related bacterial infections.

“What this study adds?

We find trends suggesting that rates of nonfatal overdose and injection-related bacterial infections decline during pregnancy and then rebound postpartum. These findings, if confirmed in future research, suggest a clear path toward intervention development: partnering with people who use drugs to design interventions that complement and support their existing harm reduction interventions during pregnancy and in the postpartum period.

## Introduction

Pregnant and postpartum people who use drugs live at the intersection of two of the gravest public health threats confronting the 21^st^ century US: crises of (1) maternal morbidity and mortality, and (2) drug-related harms. Overdoses, substance use disorders, and other mental health conditions are now among the leading causes of pregnancy-related deaths in most states,(1) a staggering fact in a nation with the highest maternal mortality rate of all high-income countries.(2) Public health has, however, only recently mobilized to end crises of maternal drug-related harms.(3) This complacency may have roots in part in complementary forms of neglect within the fields of maternal health and harm reduction: maternal health has long ignored behavioral health, including drug-related harms,(3, 4) and harm reduction has long neglected the health of women in general and of pregnant people specifically.(5) As a result, related evidence about distributions and determinants of overdoses and other drug-related harms among pregnant/postpartum people remains nascent. This evidence is critical to developing, implementing, and evaluating policy and programmatic interventions designed to help people who use drugs (PWUD) survive the 21 months they are pregnant and postpartum.

The purpose of the present analysis is to build this essential evidence, with a focus on two sets of understudied but serious drug-related health outcomes: (1) nonfatal overdoses, and (2) injection-related bacterial infections. To date, all research on maternal overdoses has analyzed fatal overdoses. *Nonfatal* overdoses, however, are important health outcomes in and of themselves: in the general (nonpregnant) population, nonfatal overdoses can cause significant neurological and cardiac complications(6, 7) Among pregnant people, nonfatal overdoses can also precipitate miscarriage and stillbirth.(8) Moreover, nonfatal overdoses are strong predictors of future fatal overdoses,(9) and thus are essential intervention points.

Research with the general (nonpregnant) population testifies to the importance of analyzing the burden of injection-related bacterial infections among pregnant/ postpartum PWUD. Injecting with unsterile syringes, cookers, cotton, or unsterile water – or injecting into unsterile skin – introduces bacteria into the skin and circulatory system that cause serious invasive bacterial infections.(10) Psychobehavioral changes may make these infections more common during pregnancy. Syringe service programs, for example, reduce the risk of injection-related bacterial infections by distributing sterile syringes and other injection equipment, but visibly pregnant PWUD report avoiding these programs to reduce stigma.(11–17) Two injection-related bacterial infections hold particular importance because of their severity: infective endocarditis (hereafter, “endocarditis”) and abscesses/cellulitis. Endocarditis is a bacterial infection of the heart that is fatal if untreated. Rates of injection-related endocarditis hospitalizations in the general population almost tripled between 1999-2017.(18–20) Meta-analyses of hospital-based case series data find that 73%-100% of ***maternal*** endocarditis cases are injection related;(21, 22) and that endocarditis’ fatality rate during pregnancy is 17.2%.(22)

One-third to one-half of all PWUD who inject in the general population report a recent abscess or cellulitis.(23–25) Abscesses are pockets of pus that typically occur at the injection site when bacteria are introduced subcutaneously; cellulitis is a painful skin infection that can cause bacteremia, suppurative arthritis, and endocarditis. These infections are the most common cause of PWUD hospitalization,(26) and have a fatality rate of 5% among hospitalized PWUD.(27)

To support the development of interventions to help PWUD survive the 21 months of pregnancy and the postpartum period, the present analysis describes, for the first time, population-based rates of hospital encounters for (1) nonfatal overdose and (2) three injection-related bacterial infections – namely, endocarditis, abscesses, and cellulitis – across the 21 months of pregnancy and the postpartum period in one state (New York State). Inequities in these outcomes exist in the general population across groups defined by race/ethnicity and other dimensions of social position.(28) Moreover, research on other maternal drug-related harms suggests that they are dynamic across the 21 months of pregnancy and the postpartum period.(29) To support tailored interventions, we thus analyze overdoses and injection-related bacterial infections by social position, and by pregnancy trimester and postpartum quarter.

## Methods

### Overview and data source

This is a retrospective cohort study of a census of individuals who experienced an in-hospital delivery in NYS between 2016-2018. We analyzed hospital discharge data (HDD) on all hospital encounters (emergency department, inpatient, and outpatient/ambulatory) regardless of payor within the state; HDD were acquired via the Healthcare Cost and Utilization Project (HCUP). We focus on NYS because (1) it is one of a subset of states with HDD that includes a unique patient identifier linking encounter records made by the same person across time and facilities, and (2) during the study period, NYS had an overall maternal mortality rate per live births close to the overall rate for the US (NYS:20.8; US:17.4).(30)

### Cohort Formation

The index event to enter the cohort was an in-hospital delivery in a NYS hospital between 9/1/2016 and 1/1/2018; live births and fetal deaths were included. We selected this window to capture all hospital encounters occurring during the presumed 9 months of pregnancy (i.e., back to 1/1/2016 for individuals who delivered on 9/1/2016) and throughout the postpartum year (i.e., up through 12/31/2018 for individuals who delivered 1/1/2018) for all cohort members. Deliveries were identified using the Alliance For Innovation On Maternal Health (AIM) Birth Admission Code List algorithm, which analyzes diagnostic fields, procedure fields, and DRGs. This algorithm includes stillbirths and excludes spontaneous and induced abortion and ectopic pregnancies. When a patient had >1 delivery during the study period, we use the most recent delivery as the index event.

## Measures

### Prevalence of Nonfatal Overdose

We created two sets of nonfatal overdose prevalence measures, each capturing a different temporal window: a prevalence estimate spanning the 21 months of pregnancy and the postpartum year, and prevalence estimates for each pregnancy trimester and postpartum quarter; we also calculated prevalence estimates for the delivery visit itself, and for the delivery month excluding that visit. The denominator for each prevalence estimate was the total number of deliveries in the group of interest during the analytic period (e.g., prevalence estimates for White people used the total number of White people who delivered within study period as the denominator). Across all measures, the prevalence numerator was the total number of cohort members in the group of interest diagnosed a nonfatal overdose in at least one hospital encounter during the analytic time period of interest; individuals who had >1 encounter for the same type of harm during a single analytic time period contributed one case to the numerator. We assessed pregnancy trimesters and postpartum quarters using the delivery date as the anchor; pregnancy trimester estimates assumed a full-term pregnancy. Box 1 depicts the diagnoses of interest and associated ICD-10 codes for overdoses from opioids, psychostimulants, sedatives/hypnotics, hallucinogens, and cannabis; following best practices,(31) overdose diagnoses were drawn from any one of the NYS HDD’s 35 diagnostic fields. Given this paper’s focus, we excluded overdoses that ended in death prior to discharge.

Jeffrey’s confidence limits where computed for the overdose prevalence measures, utilizing intervals based on the noninformative Jeffreys prior for the binomial proportion.(32) This method appropriately sets the lower limit to zero and avoids negative lower limits that can occur for rare events.

#### Box 1. Diagnoses of interest and associated ICD-10 codes

##### Overdose

T40.1 (heroin), T40.2 (other opioids), T40.3 (methadone), T40.4 (synthetic narcotics), T40.5 (cocaine), T40.6 (unspecified narcotics), T42.3 (barbiturates), T42.4 (benzodiazepines), T42.6 (sedative-hypnotics), T42.7 (unspecified sedative-hypnotics), T43.60 (psychostimulants), T43.62 (amphetamines), T43.63 (methylphenidate), T43.64 (ecstasy), T43.69 (other psychostimulants), T40.7 (cannabis), T40. (lysergide), T40.9 (psychodysleptics)

##### Injection-related bacterial infections

***Infective Endocarditis:*** B37.6 (Candidal endocarditis), I33.0 (Acute and subacute infective endocarditis), I33.9 (Acute and subacute endocarditis, unspecified), I38 (Endocarditis, valve unspecified), I39 (Endocarditis and heart valve disorders in diseases classified elsewhere)

***Abscess/cellulitis:*** L02.1 (abscess of the neck), L02.4 (abscess of the limb), L02.5 (abscess of the hand), L02.6 (abscess of the foot); L03.111 (cellulitis of the right axilla); L03.112: cellulitis of the left axilla; L03.113 cellulitis of the right upper limb; L03.114: cellulitis of the left upper limb; L03.115: cellulitis of the right lower limb; L03.116 (cellulitis of the left lower limb), L03.119 (cellulitis, unspecified limb), L03.221 (cellulitis of the neck), L03.314 (cellulitis of the groin)

### Prevalence of injection-related bacterial infections

These measures were calculated as above, except for the diagnostic codes that contributed to the numerator (Box 1). To ascertain cases of injection-related endocarditis, we used Ball et al’s roster of ICD codes.(33) Because hospital-specific case series analyses indicate that 73%-100% of all individuals who are diagnosed with infective endocarditis during pregnancy or the postpartum period are living with *injection-related* endocarditis, we did not exclude patients based on competing causes (e.g., congenital heart disease).

ICD codes specify infection location. Because abscesses and cellulitis overwhelmingly occur at the injection site, (34) we selected codes to ascertain these infections that specify sites where people commonly inject (e.g., arms, groin, feet).(35, 36) Confidence intervals were calculated as above.

### Sociodemographic characteristics

We analyzed HDD data from the delivery visit to characterize patient race/ethnicity, rurality, insurance payor at delivery, and age at delivery. Rurality was assessed by categorizing patient’s county of residence at delivery using USDA RUCC codes. If data were missing at delivery, we used data from the most temporally proximate encounter. Because of small cell sizes, we collapsed some categories (e.g., Medicaid and Medicare).

### Relative Prevalence Ratio

Prevalence ratios covering the 21 months of pregnancy and the postpartum period and their corresponding confidence intervals were computed comparing subpopulations using the SAS NLmeans macro. We derived standard errors on the logistic scale to create confidence intervals since the prevalences are small.

### Prevalence Difference

Prevalence differences covering the 21 months of pregnancy and the postpartum period were estimated comparing subpopulations by subtracting the prevalence in Group *i* from the prevalence in the reference group. Confidence intervals were computed using the standard error for differences in proportions.(37)

Reference groups for prevalence differences and prevalence ratios were selected based on two criteria: (1) prevalence and (2) case counts. We sought the group with the lowest prevalence that had a large enough case count to support robust inter-group comparisons (i.e., Black people for racial/ethnic comparisons, ages 30-34 for age comparisons, privately insured for insurance status, and large metro areas for rurality comparisons).

Data were processed and analyzed using SAS V9.4 (SAS Institute, Inc., Cary, NC, USA). Code can be found at [insert Github link after accepted].

Research was conducted consistent with ethical principles and approved by the authors’ Institutional Review Board.

## Results

### Sample description

Between 9/1/2016 and 1/1/2018, 330,872 people delivered in an NYS hospital (Table 1). The modal age category was 30-34 at delivery. Approximately half the sample (47.9%) identified as non-Hispanic White, 14.8% were non-Hispanic Black/African American, 17.1% were Hispanic/Latinx. Around half (51.0%) of the delivery visits were covered by Medicare/Medicaid; almost all publicly insured people were on Medicaid. The majority of birthing parents (82.5%) resided in a large metropolitan area at delivery, 12% resided in a small metropolitan area, 5.5% lived in either a micropolitan area or rural area.

**Table 1-.** Sociodemographic characteristic at delivery among a census of people who delivered in New York State between 9/1/2016 and 1/1/2018.

| Characteristic | Total |
| --- | --- |
| Total sample, *N* | 330,872 |
| Race/ethnicity, *n* (%) |  |
| White | 158,593 (47.9%) |
| Black/African-American | 48,923 (14.8%) |
| Hispanic/Latinx | 56,667 (17.1%) |
| Other | 66,689 (20.2%) |
| Age, *n*(%) |  |
| 10-24 | 61,922 (18.7%) |
| 25-29 | 86,050 (26.0%) |
| 30-34 | 103,366 (31.2%) |
| 35+ | 79,526 (24.0%) |
| Missing | 8 (0.0%) |
| Insurance Status, *n* (%) |  |
| Medicare/Medicaid | 168,734 (51.0%) |
| Private Insurance | 155,708 (47.1%) |
| Other | 6,355 (1.9%) |
| Missing | 75 (0.0%) |
| Rurality, *n* (%) |  |
| Large Metro Area | 272,806 (82.5%) |
| Small Metro Area | 39,656 (12.0%) |
| Non-micro/Rural | 18,324 (5.5%) |
| Missing | 86 (0.0%) |

#### Patterns of nonfatal overdose prevalence

Data indicate that 158 people per 100,000 births (CI: 145/100,000, 172/100,000; Table 2) were diagnosed with a nonfatal overdose at least once during the 21 months they were pregnant/ postpartum. While confidence intervals by trimesters and postpartum quarters overlapped, there was a trend suggesting that nonfatal overdose prevalence declined across the three pregnancy trimesters, from a high of 20/100,000 (CI: 16/100,000, 26/100,000) in the first trimester to a low of 15/100,000 (CI: 11/100,000, 19/100,000) in the third. The rate of nonfatal overdose was exceptionally high at the delivery visit itself: 41/100,000 people experienced a nonfatal overdose during these typically 3-4 day stays (CI: 34/100,000, 48/100,000). Rates were low for all other days during the delivery month (7/100,000; CI: 5/100,000, 10/100,000). Overdose rates returned to approximately their first trimester high in the first postpartum quarter (22/100,000; CI: 17/100,000, 27/100,000), and that rate was sustained across the final three postpartum quarters.

**Table 2-.**
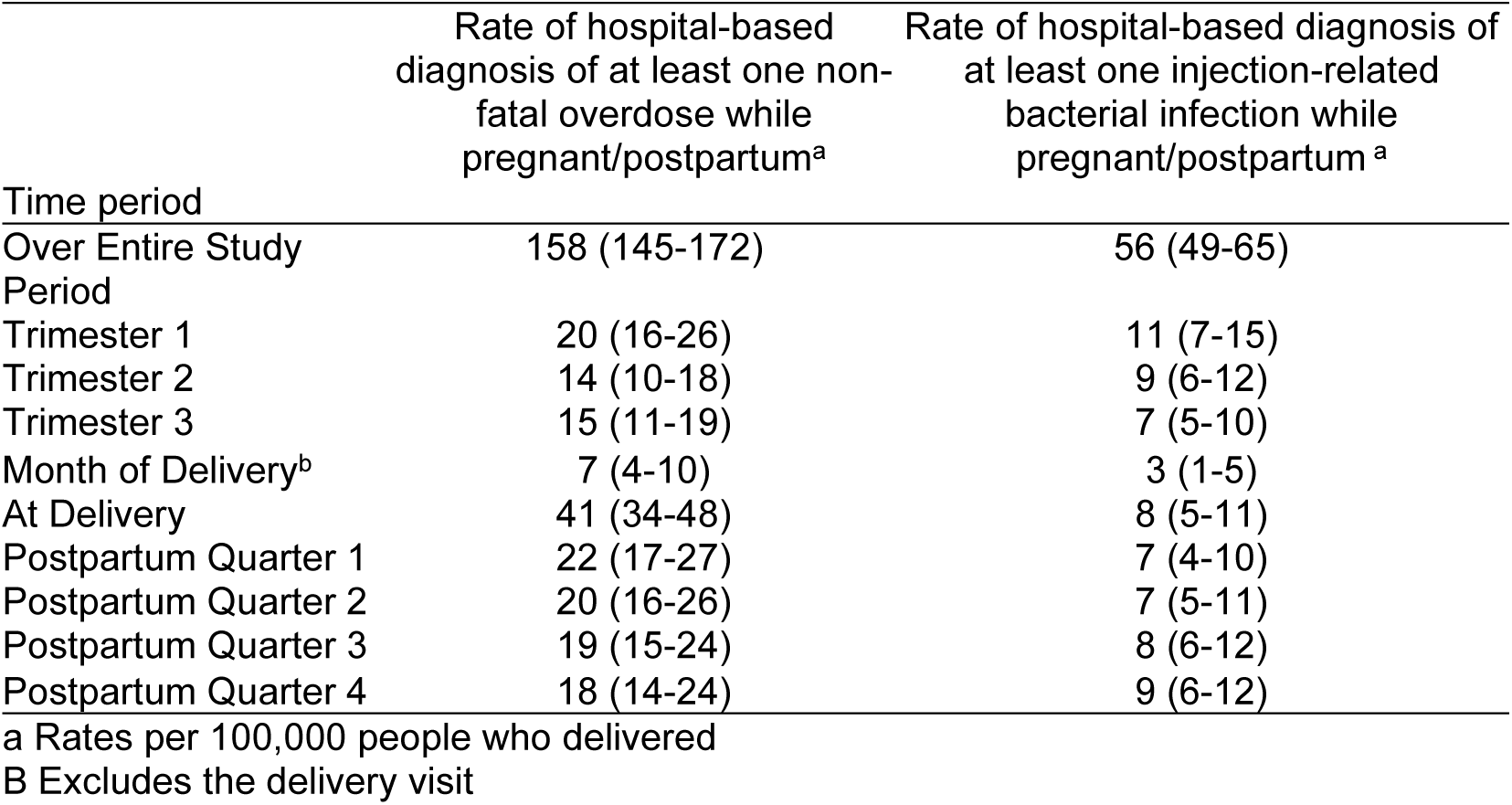
Rates of hospital-based diagnosis of non-fatal overdose and injection-related bacterial infections per 100,000 people who delivered in New York State between 9/1/2016 and 1/1/2018 (N=330,872), by pregnancy period.

| Time period | Rate of hospital-based diagnosis of at least one non-fatal overdose while pregnant/postpartum<sup>a</sup> | Rate of hospital-based diagnosis of at least one injection-related bacterial infection while pregnant/postpartum <sup>a</sup> |
| --- | --- | --- |
| Over Entire Study Period | 158 (145-172) | 56 (49-65) |
| Trimester 1 | 20 (16-26) | 11 (7-15) |
| Trimester 2 | 14 (10-18) | 9 (6-12) |
| Trimester 3 | 15 (11-19) | 7 (5-10) |
| Month of Delivery<sup>b</sup> | 7 (4-10) | 3 (1-5) |
| At Delivery | 41 (34-48) | 8 (5-11) |
| Postpartum Quarter 1 | 22 (17-27) | 7 (4-10) |
| Postpartum Quarter 2 | 20 (16-26) | 7 (5-11) |
| Postpartum Quarter 3 | 19 (15-24) | 8 (6-12) |
| Postpartum Quarter 4 | 18 (14-24) | 9 (6-12) |

a Rates per 100,000 people who delivered
B Excludes the delivery visit

Nonfatal overdose rates across the 21 months varied by social position (Table 3), generating relative prevalence ratios and prevalence differences of considerable magnitude (Table 4). While prevalence ratios found no significant White/Black comparison of overdose rates (1.23, CI: 0.94, 1.51), we find that people who identified as Latinx or any “other” race had lower overdose rates than their Black counterparts (Latinx vs. Black: 0.49, CI: 0.32, 0.66; Other vs. Black: 0.30, CI: 0.18, 0.42); prevalence differences followed a similar pattern. Prevalence ratios indicate that people aged 10-24 or 25-29 had overdose rates 68-70% higher than the reference group of people aged 30-34 (10-24: 1.70, CI: 1.27-2.12; 25-29: 1.68 (1.29-2.07); rate ratios for those over 35 years old were statistically indistinguishable from the reference group. Prevalence differences suggest that people slightly younger (25-29 years old) than the modal age of group of 30-34 years suffered approximately 81/100,000 more overdoses over the study period. People on public insurance (i.e., Medicare or Medicaid) had nonfatal overdose rates nearly three times higher than their privately insured counterparts (Medicare/Medicaid vs. Private: 2.78, CI: 2.23, 3.33), suffering excess cases of 149/100,000 (CI: 122/100,000, 176/100,000). Residents of small metro areas and micropolitan areas/rural areas had nonfatal overdose rates 2-2.5 times greater than residents of large metro areas, and suffered between 145/100,000 and 192/100,000 excess cases (Table 4).

**Table 3-.** Prevalence of hospital-based diagnoses of non-fatal overdoses and injection-related bacterial infections per 100,000 while pregnant/postpartum among people who delivered in New York State between 9/1/2016 and 1/1/2018 (N=330,872), by social position.

| Characteristic | Prevalence<sup>a</sup> of hospital-based diagnosis of at least one non-fatal overdose while pregnant/postpartum (95% CI) | Prevalence<sup>a</sup> of hospital-based diagnosis of at least one injection-related bacterial infection while pregnant/ postpartum (95% CI) |
| --- | --- | --- |
| Total sample | 158 (145-172) | 56 (49-65) |
| Race/ethnicity |  |  |
| White | 221 (198-245) | 90 (76-105) |
| Black/African-American | 180 (145-220) | 43 (27-64) |
| Hispanic/Latinx | 88 (66-115) | 30 (18-47) |
| Other | 54 (38-74) | 9 (4-19) |
| Age |  |  |
| 10-24 | 202 (169-240) | 63 (45-85) |
| 25-29 | 200 (172-231) | 80 (63-101) |
| 30-34 | 119 (99-141) | 45 (33-59) |
| 35+ | 131 (107-158) | 40 (28-56) |
| Insurance Status |  |  |
| Medicare/Medicaid | 232 (210-256) | 100 (85-115) |
| Private Insurance | 83 (70-99) | 11 (7-17) |
| Other | 31 (7-101) | 16 (2-74) |
| Rurality |  |  |
| Large Metro Area | 130 (117-144) | 45 (38-54) |
| Small Metro Area | 275 (227-330) | 108 (80-145) |
| Non-micro/Rural | 322 (248-412) | 98 (60-152) |

a Rates per 100,000 mothers who delivered

**Table 4-.** Disparities in the prevalences of hospital-based diagnoses of non-fatal overdoses and injection-related bacterial infections per 100,000 while pregnant/postpartum among people who delivered in New York State between 9/1/2016 and 1/1/2018 (N=330,872)

| Characteristic | Non-Fatal Overdoses (95% CI) |  | Injection-related Bacterial Infections (95% CI) |  |
| --- | --- | --- | --- | --- |
|  | Relative Risk Ratios<sup>a</sup> | Difference of Rates<sup>a</sup> | Relative Risk Ratios<sup>a</sup> | Difference of Rates<sup>a</sup> |
| Total sample | 158 | 158 | 56 | 56 |
| Race/ethnicity (ref: Black/African-American) |  |  |  |  |
| White vs. Black | 1.23 (0.94-1.51) | 41 (-3-85) | 2.09 (1.13-3.04) | 47 (23-70) |
| Hispanic vs. Black | 0.49 (0.32-0.66) | -92 (-136--47) | 0.7 (0.25-1.15) | -13 (-36-10) |
| Other vs. Black | 0.3 (0.18-0.42) | -126 (-167--84) | 0.21 (0.02-0.4) | -34 (-54--14) |
| Age (ref: 25-29) |  |  |  |  |
| 10-19 vs. 25-29 | 1.7 (1.27-2.12) | 83 (42-124) | 1.42 (0.81-2.02) | 18 (-5-42) |
| 20-24 vs. 25-29 | 1.68 (1.29-2.07) | 81 (44-117) | 1.8 (1.13-2.47) | 36 (13-59) |
| 30-34 vs. 25-29 | 1.1 (0.81-1.39) | 12 (-21-45) | 0.9 (0.5-1.31) | -4 (-23-15) |
| Insurance Status (ref: Private) |  |  |  |  |
| Public vs. Private | 2.78 (2.23-3.33) | 149 (122-176) | 9.11 (4.57-13.66) | 89 (73-105) |
| Rurality (ref: Large metro area) |  |  |  |  |
| Small vs. Large Metro Area | 2.11 (1.66-2.57) | 145 (91-198) | 2.39 (1.56-3.21) | 63 (30-96) |
| Rural/micro vs Large Metro | 2.47 (1.79-3.16) | 192 (109-275) | 2.16 (1.09-3.23) | 53 (7-99) |

a Rates per 100,000 people who delivered

#### Patterns of injection-related bacterial infection prevalence

Across these 21 months, 56 people for every 100,000 people who delivered (CI: 49/100,000, 65/100,000) were diagnosed at least once with an injection-related bacterial infection (Table 2). The prevalence of these infections was relatively constant across pregnancy trimesters and postpartum quarters, hovering between 7-11 cases/100,000. Notably, the rate during the delivery visit itself was 8/100,000 (CI: 1/100,000, 5/100,000), a striking figure for the typical 3-4 day hospital stay at delivery that is comparable to rate that spanned entire trimesters/quarters.

Prevalence varied by social position (Tables 3 and 4). Prevalence ratios indicated that White people appeared to have around twice the rate of injection-related bacterial infections (2.09, CI: 1.13, 3.04) relative to their Black counterparts. Risk differences indicate that members of the “Other” racial/ethnic group experienced 34 fewer cases per 100/000 compared to their Black counterparts (CI: −54/100,000, −14/100,000). As with non-fatal overdoses, younger people suffered more: people aged 25-29 years old had nearly double the rate of the modal age group (1.80; CI: 1.13, 2.47), and people aged 10-24 had nearly 18 more cases (CI: −5/100,000, −142/100,000). People on Medicare/Medicaid had 9 times the rate of these infections than privately insured people (9.11, CI: 4.57, 13.66), and 89 excess infections (CI: 73, 105). People living outside large metro areas had 2.2-2.4 times the prevalence of these infections compared to residents of large metro areas, and 53-63 excess cases.

## Discussion

Analyses of HDD data from NYS indicate that the prevalence of maternal hospital encounters for nonfatal overdose diagnoses was 158 cases per 100,000 people who delivered across the 21 months of pregnancy and the postpartum period. Rates of injection-related bacterial infections across these 21 months were approximately 1/3 that rate, 56/100,000. Endocarditis, however, has a high fatality rate during pregnancy,(22) and the long hospital stays often required to treat endocarditis, abscesses, and cellulitis may seriously disrupt the families, workplaces, and broader communities in which these patients are embedded and incur significant cost.(38, 39)

Risk ratios and risk differences reveal a pattern of striking inequities by rurality/ urbanicity and payor: residents living outside of large metropolitan areas suffered far higher rates of the outcomes of interest than residents living inside these areas, and correspondingly suffered considerable excess cases per 100,000. A similar pattern emerged for people insured by public programs (i.e., Medicaid and Medicare, though in this sample these patients were overwhelmingly covered by Medicaid) compared to their privately insured counterparts, and both parallel findings from this same cohort on substance use disorder diagnoses.(40) Access to harm reduction programs may drive these inequities: large metropolitan areas are home to far higher densities of substance use disorder treatment programs(41) and syringe service programs(42) than other US regions, and a high percentage of treatment programs refuse to accept Medicaid.(43) In the US, healthcare coverage is entangled with economic status, and so the high rates among publicly insured individuals may also reflect poverty’s toll above and beyond health insurance, including persistent ill-health and physiological stress that create vulnerability to overdose and bacterial infections.(44, 45)

Racial/ethnic disparities were complex, but broadly follow documented patterns of substance use disorders among pregnant people. Parallel to our results, analyses of data from the National Survey on Drug Use and Health indicate that Black and White pregnant people have similar rates of substance use disorder diagnosis.(46) Note, however, that data from more recent years might reveal a different pattern: the prevalence of overdoses among Black people in the general (nonpregnant) population surpassed the prevalence among White people in the late 2010s.(47)

There was a trend suggesting a decline in nonfatal overdoses and injection-related bacterial infections across the three trimesters of pregnancy, followed by a postpartum rebound. The decline is consistent with qualitative research, which has found that PWUD attempt to reduce or cease using drugs when they learn they are pregnant, to protect their own health and that of their neonate.(15, 16, 48, 49) If the decline is confirmed in future research with larger sample sizes, it may testify, in part, to the determination of pregnant people to protect their own health and that of their neonate, often in the face of multiple pregnancy-related challenges (e.g., few substance use disorder treatment programs accept pregnant people(50)). Ascertainment bias may, however, amplify this pattern of decline and rebound: PWUD may be increasingly reluctant to seek care for drug-related harms as their pregnancies become visible, and this reluctance may dissipate postpartum.

If confirmed in future research, a postpartum rebound marks a crucial opportunity to support PWUD who reduced or ceased using drugs or injecting while pregnant, and wish to sustain this change postpartum. Interventions include buprenorphine induction at delivery, mobile harm reduction programs that offer at-home delivery of harm reduction materials, and interweaving harm reduction efforts into postpartum home visiting programs. Most notably, seven states have enacted laws prioritizing mothers of minor children for substance use disorder treatment, highlighting a key policy opportunity for the remaining 43 states to support PWUD postpartum.

Rates of all nonfatal overdose and of injection-related bacterial infections were strikingly high at the delivery visit, and a pattern that might reflect provider response to these harms. Providers treating a patient who is overdosing or experiencing endocarditis and is late in their pregnancy may advise the patient to induce labor or perform a c-section to protect maternal and fetal health.(51) Additionally, patients admitted for delivery may choose to simultaneously seek care for an abscess/ cellulitis; delivery visits may also offer providers opportunities to detect these infections.

### Limitations

Data were derived from a single state, NYS, which did not require healthcare providers to report drug use in pregnancy to child protective services during the study period. Patterns and inequities in drug-related harms might differ in states that mandate reporting. Patterns and inequities might also differ by state Medicaid environment: NYS had expanded Medicaid through the full postpartum year by the onset of the study period. Our study period ended before the COVID pandemic, and subsequent years witnessed surging fatal overdose rates across multiple populations and shifting inequities in overdoses.(40) Future research should explore whether and how the patterns found here changed during and after the pandemic. While we employed validated algorithms to identify cases of overdose and endocarditis,(31, 33) we relied on injection sites to ascertain cases of abscess/cellulitis. Injection locations, may, however, shift during pregnancy, as pregnant PWUD seek to manage stigma by concealing ongoing drug use.

## Conclusions

If confirmed in future research with larger sample sizes, findings hold promise for future interventions. The trend toward declining rates of diagnoses for nonfatal overdose and injection-related bacterial infections during pregnancy, combined with extant qualitative research, suggest that PWUD engage in harm reduction efforts while pregnant. Public health should partner with PWUD to understand these efforts, and to develop interventions that complement and support them. Subsequent rebounds in these rates testify to the urgent need to co-develop interventions with PWUD that support the persistence and adaptation of these harm reduction efforts postpartum.

Interventions should be tailored to highly vulnerable populations (e.g., rural areas) to ensure that they do not exacerbate existing inequities.

## Data Availability

All data were obtained through the Hospital Cost and Utilization Project (HCUP). To access those data, please visit https://hcup-us.ahrq.gov/

## Acknowledgements

We are grateful to the Hospital Cost and Utilization Project for creating the database. We are also grateful to the following funding sources: [add post review].

## Declarations

All authors have approved this manuscript.

## Funding

Wien and Rogers were supported by T32DA050552. Cooper, Blake, and Kramer were supported by U54HD113292; Cooper was also supported by the following NIH grants: R49CE003072; R01DA058065; R01DA046197.

## Conflicts of interest/Competing interests

We have no conflicts to disclose.

## Ethics approval

The Emory Institutional Review Board approved all analyses.

## Consent to participate

N/A. Analysis of existing hospital discharge data.

## Consent for publication

N/A. Analysis of existing hospital discharge data.

## Authors’ contributions

All authors guided the analyses, supported the interpretation of findings, and reviewed and edited the manuscript. In addition, Cooper drafted the original manuscript, and D’Souza conducted analyses and drafted tables.

## Notes

### Competing Interest Statement

The authors have declared no competing interest.

### Author Declarations

The Institutional Review Board at Emory University gave ethical approval for this work.

